# Prediction of Long COVID and Mortality among Patients with Substance Use Disorder

**DOI:** 10.1101/2025.11.19.25340587

**Authors:** Jiawei Wu, K M Sajjadul Islam, Praveen Madiraju

## Abstract

The convergence of the COVID-19 pandemic and the substance use disorder (SUD) crisis has created a syndemic that places this vulnerable population at extreme risk for acute and chronic adverse outcomes. This study addresses the critical need for proactive risk stratification by developing and contrasting machine learning models to predict two distinct endpoints in hospitalized patients with COVID-19 with SUD: in-hospital mortality and long COVID. Using comprehensive electronic health record (EHR) data, we systematically address severe class imbalance using a combination of specialized algorithms (e.g., Balanced Random Forest) and data resampling techniques (e.g., SMOTE). Our fine-tuned Logistic Regression model for mortality achieves 93% recall, successfully identifying patients at risk of death. For the more challenging long COVID prediction task, our proposed weighted ensemble model achieves 80% recall, demonstrating strong performance in identifying patients susceptible to chronic illness. Feature importance analysis reveals distinct clinomic signatures: acute mortality is driven by markers of systemic distress (e.g., lactic acid, D-dimer), while chronic risk is linked to metabolic and inflammatory factors (e.g., BMI, renal function, preexisting sleep disorders). Our work delivers a validated computational toolkit for dual-risk prediction, enabling targeted interventions to mitigate both immediate and long-term harm in this high-risk population.

## I. Introduction

The convergence of the COVID-19 pandemic and the pre-existing public health crisis of substance use disorders (SUD) has created a devastating syndemic. Individuals with SUD are a uniquely vulnerable population, facing compounded physiological and psychosocial risks that increase their susceptibility to the full spectrum of complications of COVID-19 [1], [2]. This dual threat manifests itself as both an acute and chronic crisis. The acute danger to this population is well documented. Factors such as compromised lung function, particularly in those with Opioid Use Disorder (OUD), a higher prevalence of comorbidities, and dysregulated inflammatory responses can dramatically increase the risk of in-hospital death [3]. Chronically, persistent fatigue, cognitive deficits, and respiratory problems associated with long COVID can initiate a vicious cycle, where debilitating symptoms exacerbate substance use and co-occurring mental health disorders (MHD), severely deteriorating quality of life and complicating recovery [4]–[7]. A previous work established the statistical foundation, confirming that COVID-19 patients with SUD face significantly higher odds of severe outcomes and demonstrating the utility of machine learning to predict length of stay [8]. However, that foundational analysis did not build dedicated predictive models for the two most clinically critical endpoints: mortality and long COVID. The former requires identifying patients at immediate risk of decompensation; the latter requires identifying those susceptible to long-term illness. These are distinct predictive tasks with potentially different biological drivers that require separate, customized models.

This paper directly addresses this gap. We develop, validate and compare machine learning models to predict both in-hospital mortality and the onset of long COVID in patients with SUD. Our primary contributions are listed below:

1. Develop and validate a robust machine learning model to predict in-hospital mortality among COVID-19 patients with SUD.
2. Develop and validate a model to predict the onset of long COVID in this population and analyze its strong association with co-occurring mental health disorders.
3. Identify and contrast the key clinical features that drive these distinct acute versus chronic outcomes, providing insights into their potentially different underlying mechanisms.

By delivering a validated toolkit for dual risk prediction, this work provides a more holistic approach to management of COVID-19 in patients with SUD and enables interventions that can mitigate both immediate and long-term harm.

## II. Related Work

### A. The Clinical Landscape: SUD as a Risk Amplifier for COVID-19

A substantial body of research confirms that individuals with SUD experience disproportionately severe COVID-19 outcomes. Foundational studies using large-scale EHRs established SUD, particularly Opioid Use Disorder (OUD), as a significant independent risk factor for infection, hospitalization, and mortality [3], [9]. These analyses primarily employed statistical methods, such as multivariable logistic regression, to calculate adjusted odds ratios (aORs) and identify associated risk factors. While crucial for establishing clinical risk, these explanatory models are not designed for patient-level prediction. Concurrently, the syndemic of long COVID, SUD, and co-occurring mental health disorders (MHD) has been described, highlighting a self-perpetuating cycle of physical and psychological distress [10], [11]

### B. The Methodological Shift: From Statistical Association to Clinical Prediction

In parallel, machine learning (ML) has been successfully applied to forecast COVID-19 outcomes, identifying complex patterns in high-dimensional data that regression models may miss [12], [13]. Several studies have specifically applied ML to predict long COVID in the general population. For example, Purohit et al. successfully used ML to predict the onset of MHD following a long COVID diagnosis, achieving an AUC of above 0.90 [14], while Patterson et al. used ML with cytokine biomarkers to differentiate long COVID from other chronic illnesses [15]. These studies demonstrate the power of ML to generate predictive, clinically relevant insights.

### C. The Research Gap and Our Contribution

Despite these advances, a critical research gap persists. COVID-19 research in the SUD population has largely been associational, describing risk rather than forecasting it. In contrast, predictive ML models for long COVID have focused on the general population, overlooking the specific complexities of SUD patients.

To our knowledge, no previous study has developed and applied predictive models. This study addresses a literature gap by developing and applying predictive models, including tree-based models and the Tab-Transformer deep learning architecture [16], to predict both acute (mortality) and chronic (long-term COVID) outcomes in a high-risk cohort.

## III. Methods

Our methodology integrates clinical data processing, a multi-faceted machine learning strategy to handle severe class imbalance, and statistical analysis to uncover associations between substance use and post-acute sequelae of COVID-19.

### A. Cohort Construction and Outcome Definition

We used a de-identified EHR database from Froedtert & The Medical College of Wisconsin (Milwaukee, WI, USA), including 71,716 confirmed COVID-19 cases from March 2020 to January 2023. From all patients with a confirmed COVID-19 diagnosis, we identified a base population with a co-occurring Substance Use Disorder (SUD), defined by the presence of one or more ICD-10 codes listed in Table I. From this base population, we constructed two distinct cohorts for our predictive tasks.

**TABLE I:** ICD codes and descriptions.

| Category | ICD Codes |
| --- | --- |
| long COVID | U09.9 |
| **Substance Use Disorder (SUD)** |  |
| Alcohol Use Disorder | F10 |
| Opioid Use Disorder | F11, T40 |
| Cannabis Use Disorder | F12 |
| Sedative, Hypnotic, or Anxiolytic Use Disorder | F13 |
| Cocaine Use Disorder | F14 |
| Other Stimulant Use Disorder | F15 |
| Hallucinogen Use Disorder | F16 |
| Tobacco Use Disorder | F17 |
| Inhalant Use Disorder | F18 |
| Other (or Unknown) Substance Use Disorder | F19 |
| **Mental Health Disorder (MHD)** |  |
| Major depressive disorder, single episode | F32 |
| Major depressive disorder, recurrent episode | F33 |
| Bipolar disorder | F31 |
| Schizophrenia | F20 |
| Other anxiety disorders | F41 |
| Persistent mood disorders | F34 |
| Reaction to severe stress and adjustment disorders | F43 |
| Sleep disorders | G47 |

**TABLE II:** Patient cohort sizes for machine learning models.

| Outcome Class | Total Patients | Training (80%) | Test (20%) |
| --- | --- | --- | --- |
| In-Hospital Mortality |  |  |  |
| Survived (Negative Class) | 3,793 | 3,036 | 757 |
| Deceased (Positive Class) | 57 | 44 | 13 |
| Long COVID Diagnosis |  |  |  |
| No Long COVID (Negative Class) | 4,681 | 3,744 | 937 |
| Long COVID (Positive Class) | 138 | 111 | 27 |

#### 1) In-Hospital Mortality

3,850 hospitalized COVID-19 patients with SUD (excluding outpatient cases). The binary outcome was all-cause in-hospital mortality, identified by discharge disposition “Expired.”

#### 2) Long COVID

906 patients with long COVID (ICD-10 code U09.9), stratified into SUD-positive (n=188) and SUD-negative (n=718) subgroups (Figure 1). Co-occurring MHD was defined using ICD-10 codes (Table I), e.g., major depressive disorder (F32/F33) and anxiety disorders (F41).

**Fig. 1:**
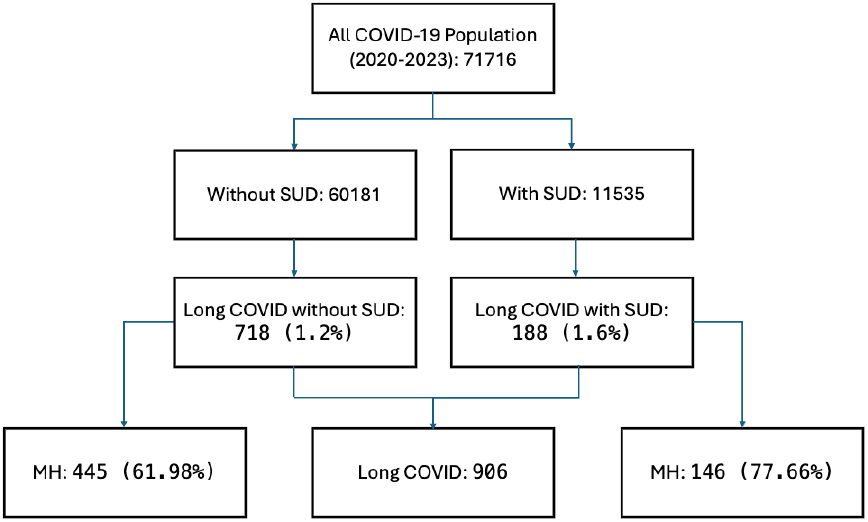
Patient selection flowchart for the long COVID analysis. The population was stratified by the presence or absence of SUD.

### B. Statistical Analysis for Long COVID and MHD Association

We used a chi-squared test to assess the statistical association between SUD and the prevalence of MHD within the long COVID patient population. This test compared the frequency of MHD diagnoses between long COVID patients with SUD and those without SUD. We performed a similar analysis to compare MHD prevalence between SUD patients who developed long COVID and those who did not.

To quantify the strength of these associations while controlling for potential confounders, we performed a multivariable logistic regression analysis to compute adjusted odds ratios (aORs) for the association between specific SUDs and a co-occurring MHD diagnosis among patients with long COVID.

### C. Predictive Modeling and Evaluation

For both mortality and long COVID prediction, we developed a pipeline designed to identify the optimal modeling strategy in a severely imbalanced setting.

#### 1) Feature Selection and Data Preparation

Our initial dataset contained a large number of features from the EHR, including demographics, encounter details, vital signs, and hundreds of laboratory results. To improve model performance, reduce computational overhead, and mitigate the risk of overfitting, we implemented a feature selection step. We employed Recursive Feature Elimination with Cross-Validation (RFECV) and identified the most informative subset of features for each task: 40 out of an initial 795 features for mortality prediction, and 79 out of 399 for long COVID prediction. The resulting cohorts were then stratified and split into 80% training and 20% holdout test sets.

#### 2) Strategy for Imbalanced Learning

Given the severe underrepresentation of the positive class in both tasks, our strategy combined data-level, algorithmic, and ensemble methods.

##### a) Baseline and Standard Models

We first established performance baselines using a suite of standard machine learning classifiers, including Logistic Regression (LR), Random Forest (RF), and XGBoost. To compare these with modern deep learning architectures, we also implemented a Tab-Transformer model as an advanced baseline. The Tab-Transformer was specifically chosen, as it is a leading deep learning architecture designed for tabular data, which utilizes self-attention mechanisms to learn contextual embeddings for categorical features and robustly map interactions between features. This makes it a strong candidate for evaluating the potential of deep learning to capture complex, non-linear patterns in EHR data compared to traditional models.

##### b) Synthetic Data Oversampling

For models not inherently designed for imbalance (LR, RF, XGBoost), we applied two oversampling techniques to the training data: the Synthetic Minority Over-sampling Technique (SMOTE) and the hybrid SMOTE with Edited Nearest Neighbors (SMOTEENN) [17].

##### c) Specialized Imbalanced Learning Algorithms

We employed two algorithms with built-in imbalance handling: Balanced Random Forest (BRF), which under-samples the majority class within each bootstrap, and Easy Ensemble (EE), which trains an ensemble of models on balanced data subsets. These methods were not combined with data-level resampling to avoid redundant balancing.

##### d) Deep Learning Imbalance Handling

For Tab-Transformer, class weights (inverse of class frequency) were applied to cross-entropy loss to penalize minority class mis-classifications.

#### 3) Advanced Ensemble Modeling for Long COVID Prediction

For the highly challenging long COVID task, we implemented an advanced ensemble method based on BRF, XGBoost and LR: a Weighted Ensemble where the final prediction is a linear combination of base model predictions, with weights optimized to maximize the recall-score on the validation set.

#### 4) Performance Evaluation

The performance of all models was systematically assessed on the test set using a comprehensive set of metrics: accuracy, F1-score, recall, precision, and the Area Under the Precision-Recall Curve (PR-AUC). Finally, a feature importance analysis was conducted on the best-performing models to identify the key clinical factors of risk for each outcome.

## IV. Results

Our results are presented in two parts. First, we establish the statistical links between Substance Use Disorder (SUD), co-occurring Mental Health Disorders (MHD), and long COVID. Second, we present the performance of our machine learning models, which were developed to predict in-hospital mortality and a long COVID diagnosis. We evaluate various models and conclude by identifying the most important clinical predictors for each outcome.

### A. Demographics and Statistical Association of Outcomes

#### a) Mortality Cohort

Analysis of the mortality research cohort (n=3,850) revealed a distinct high-risk profile (Table III). Deceased patients were significantly older (median age 63 vs. 48), more likely to be male (63.2% of deaths), and had a prolonged hospital course (mean LOS 12.4 days). Notably, specific SUDs showed divergent risk patterns: Alcohol Use Disorder was heavily overrepresented among decedents (49.1% of deaths vs. 16.7% of the entire cohort), confirming that mortality risk is heterogeneous across the SUD population and defining clear targets for predictive modeling.

**TABLE III:** Statistics for study cohort grouped by stay days and death.

| Feature | Total Population,<br>n(%) | 0-7 Day(s),<br>n(%) | 8-14 Days,<br>n(%) | 15-21 Days,<br>n(%) | 22+ Days,<br>n(%) | Death,<br>n(%) |
| --- | --- | --- | --- | --- | --- | --- |
| **Total** | 3850 (100.0) | 3374 (87.64) | 291 (7.56) | 87 (2.26) | 98 (2.55) | 57 (1.48) |
| **Age Group** |  |  |  |  |  |  |
| Median age (SD) | 48 (17) | 46 (17) | 61 (17) | 61 (14) | 60 (14) | 63 (13) |
| Mean LOS (SD) | 3.79 (5.73) | 2.08 (1.55) | 10.34 (1.96) | 17.6 (1.9) | 30.99 (7.57) | 12.37 (12.22) |
| 45-64 | 1398 (36.31) | 1203 (35.66) | 112 (38.49) | 41 (47.13) | 42 (42.86) | 25 (43.86) |
| <45 | 1685 (43.77) | 1596 (47.30) | 57 (19.59) | 13 (14.94) | 19 (19.39) | 7 (12.28) |
| >=65 | 767 (19.92) | 575 (17.04) | 122 (41.92) | 33 (37.93) | 37 (37.76) | 25 (43.86) |
| **Sex** |  |  |  |  |  |  |
| Male | 1774 (46.08) | 1483 (43.95) | 166 (57.04) | 62 (71.26) | 63 (64.29) | 36 (63.16) |
| Female | 2076 (53.92) | 1891 (56.05) | 125 (42.96) | 25 (28.74) | 35 (35.71) | 21 (36.84) |
| **Race** |  |  |  |  |  |  |
| White or Caucasian | 2166 (56.26) | 1851 (54.86) | 195 (67.01) | 57 (65.52) | 63 (64.29) | 31 (54.39) |
| Black or African American | 1454 (37.77) | 1320 (39.12) | 82 (28.18) | 25 (28.74) | 27 (27.55) | 21 (36.84) |
| American Indian or Alaska Native | 22 (0.57) | 19 (0.56) | 2 (0.69) | 1 (1.15) | 0 (0) | 1 (1.75) |
| Asian | 32 (0.83) | 28 (0.83) | 0 (0) | 1 (1.15) | 3 (3.06) | 2 (3.51) |
| Other or not specified | 176 (4.57) | 156 (4.62) | 12 (4.12) | 3 (3.45) | 5 (5.10) | 2 (3.51) |
| **Ethnicity** |  |  |  |  |  |  |
| Non Hispanic | 3637 (94.47) | 3186 (94.43) | 274 (94.16) | 83 (95.40) | 94 (95.92) | 54 (94.74) |
| Hispanic | 201 (5.22) | 177 (5.25) | 17 (5.84) | 4 (4.60) | 3 (3.06) | 3 (5.26) |
| Other or not specified | 12 (0.31) | 11 (0.33) | 0 (0) | 0 (0) | 1 (1.02) | 0 (0) |
| **Medical Problems** |  |  |  |  |  |  |
| Death | 57 (1.48) | 29 (0.86) | 12 (4.12) | 5 (5.75) | 11 (11.22) | 57 (100.00) |
| Full Vaccination | 318 (8.26) | 291 (8.62) | 20 (6.87) | 3 (3.45) | 4 (4.08) | 1 (1.75) |
| Alcohol Use Disorder | 642 (16.68) | 471 (13.96) | 98 (33.68) | 31 (35.63) | 42 (42.86) | 28 (49.12) |
| Cannabis Use Disorder | 235 (6.10) | 218 (6.46) | 12 (4.12) | 2 (2.30) | 3 (3.06) | 1 (1.75) |
| Cocaine Use Disorder | 100 (2.60) | 75 (2.22) | 13 (4.47) | 5 (5.75) | 7 (7.14) | 3 (5.26) |
| Opioid Use Disorder | 336 (8.73) | 241 (7.14) | 56 (19.24) | 17 (19.54) | 22 (22.45) | 6 (10.53) |
| Tobacco Use Disorder | 2980 (77.40) | 2714 (80.44) | 169 (58.08) | 46 (52.87) | 51 (52.04) | 28 (49.12) |

*Note.* The total population and stay-day groups already include death. We still list the death group individually for a better insight into the demographics information among the deaths.

#### b) Long COVID Cohort

Our long COVID cohort (n=906) was analyzed by stratifying patients based on a history of SUD (n=188 with SUD vs. n=718 without) (Table IV). SUD patients with long COVID were more likely to be 45–64 years old (53.19% vs. 37.33% in non-SUD) and male (43.09% vs. 33.70% in non-SUD). Black/African American patients accounted for 28.72% of SUD-long COVID cases, nearly double the non-SUD rate (14.90%). The socioeconomic is also a factor. SUD patients were 2.5-fold more likely to use Medicaid (27.66% vs. 10.86%). Additionally, SUD patients with long COVID had higher rates of MHD (The detailed statistical analysis is displayed in the next subsection).

**TABLE IV:**
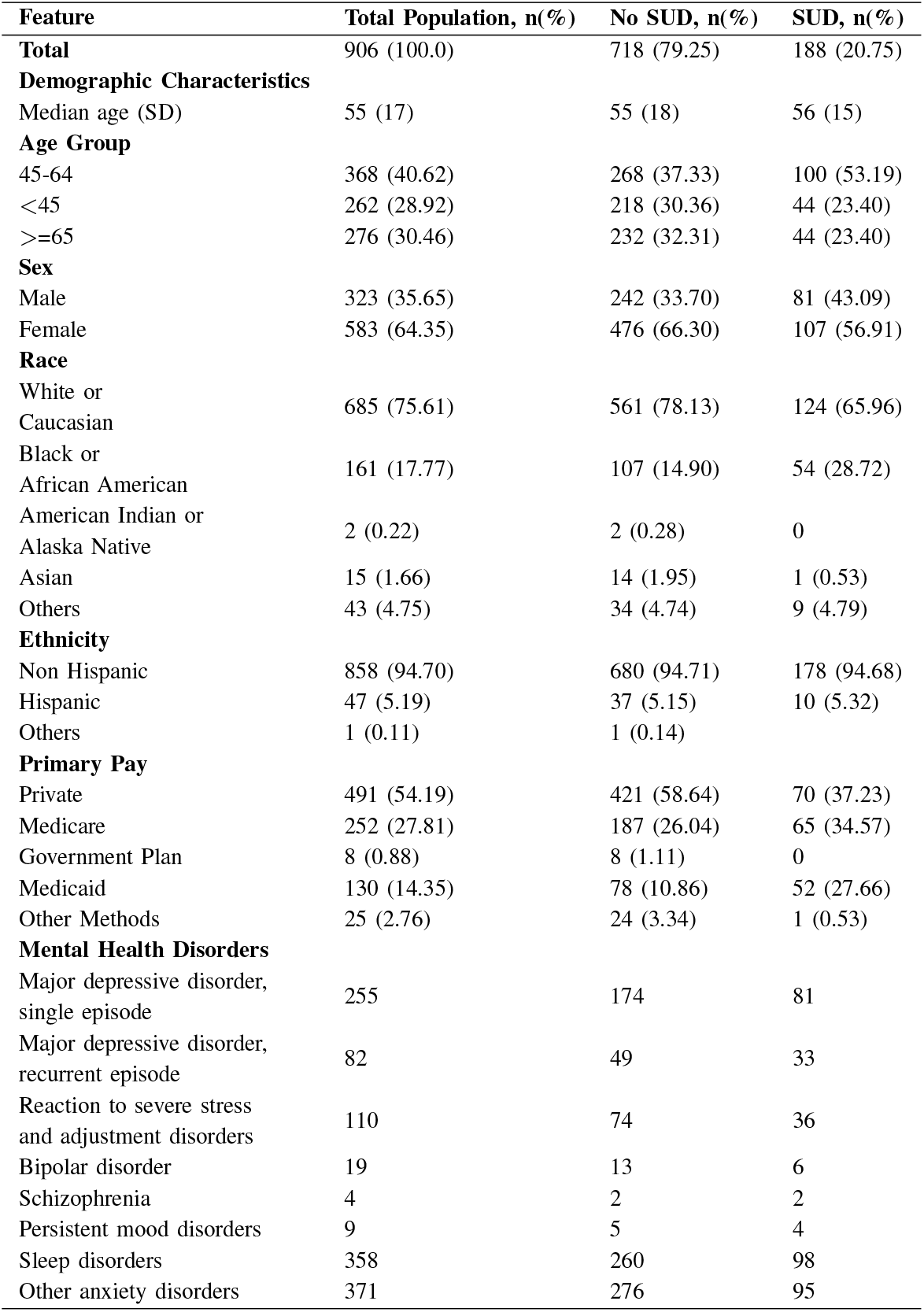
Statistics for long COVID population (SUD vs. No SUD).

| Feature | Total Population, n(%) | No SUD, n(%) | SUD, n(%) |
| --- | --- | --- | --- |
| **Total** | 906 (100.0) | 718 (79.25) | 188 (20.75) |
| **Demographic Characteristics** |  |  |  |
| Median age (SD) | 55 (17) | 55 (18) | 56 (15) |
| **Age Group** |  |  |  |
| 45-64 | 368 (40.62) | 268 (37.33) | 100 (53.19) |
| <45 | 262 (28.92) | 218 (30.36) | 44 (23.40) |
| >=65 | 276 (30.46) | 232 (32.31) | 44 (23.40) |
| **Sex** |  |  |  |
| Male | 323 (35.65) | 242 (33.70) | 81 (43.09) |
| Female | 583 (64.35) | 476 (66.30) | 107 (56.91) |
| **Race** |  |  |  |
| White or Caucasian | 685 (75.61) | 561 (78.13) | 124 (65.96) |
| Black or African American | 161 (17.77) | 107 (14.90) | 54 (28.72) |
| American Indian or Alaska Native | 2 (0.22) | 2 (0.28) | 0 |
| Asian | 15 (1.66) | 14 (1.95) | 1 (0.53) |
| Others | 43 (4.75) | 34 (4.74) | 9 (4.79) |
| **Ethnicity** |  |  |  |
| Non Hispanic | 858 (94.70) | 680 (94.71) | 178 (94.68) |
| Hispanic | 47 (5.19) | 37 (5.15) | 10 (5.32) |
| Others | 1 (0.11) | 1 (0.14) |  |
| **Primary Pay** |  |  |  |
| Private | 491 (54.19) | 421 (58.64) | 70 (37.23) |
| Medicare | 252 (27.81) | 187 (26.04) | 65 (34.57) |
| Government Plan | 8 (0.88) | 8 (1.11) | 0 |
| Medicaid | 130 (14.35) | 78 (10.86) | 52 (27.66) |
| Other Methods | 25 (2.76) | 24 (3.34) | 1 (0.53) |
| **Mental Health Disorders** |  |  |  |
| Major depressive disorder, single episode | 255 | 174 | 81 |
| Major depressive disorder, recurrent episode | 82 | 49 | 33 |
| Reaction to severe stress and adjustment disorders | 110 | 74 | 36 |
| Bipolar disorder | 19 | 13 | 6 |
| Schizophrenia | 4 | 2 | 2 |
| Persistent mood disorders | 9 | 5 | 4 |
| Sleep disorders | 358 | 260 | 98 |
| Other anxiety disorders | 371 | 276 | 95 |

### B. Statistical Results of association Between Long COVID, SUD, and MHD

The intersection of long COVID and SUD revealed a compounded mental health burden. Among patients with long COVID, those with SUD had a significantly higher prevalence of MHD than those without SUD (77.7% vs. 62.0%; p<0.001, Table V). Furthermore, the onset of long COVID within the SUD population was associated with a substantially increased prevalence of major depressive disorder (43.1% vs. 30.4%), other anxiety disorders (50.5% vs. 36.8%), and sleep disorders (52.1% vs. 27.0%) (all p<0.001, Table VI).

**TABLE V:**
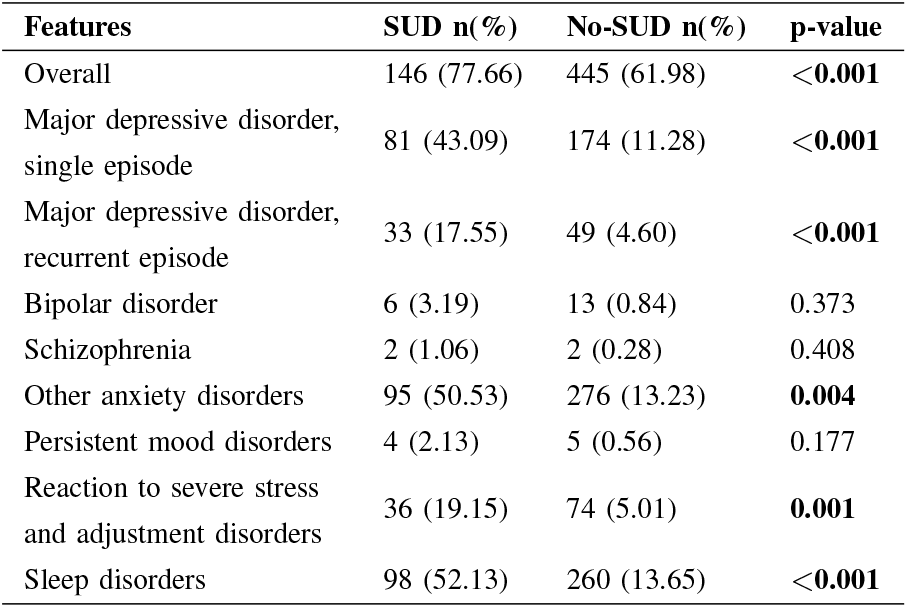
Association between SUD and MHD among long COVID patients.

**TABLE VI:** Association between long COVID (LC) and MHD among SUD patients.

| Features | LC n(%) | No LC n(%) | p-value |
| --- | --- | --- | --- |
| Overall | 146 (77.66) | 6560 (57.81) | **0.009** |
| Major depressive disorder, single episode | 81 (43.09) | 3446 (30.37) | **<0.001** |
| Major depressive disorder, recurrent episode | 33 (17.55) | 1116 (9.84) | **0.001** |
| Bipolar disorder | 6 (3.19) | 603 (5.31) | 0.260 |
| Schizophrenia | 2 (1.06) | 200 (1.76) | 0.657 |
| Other anxiety disorders | 95 (50.53) | 4178 (36.82) | **<0.001** |
| Persistent mood disorders | 4 (2.13) | 91 (0.80) | 0.112 |
| Reaction to severe stress and adjustment disorders | 36 (19.15) | 1159 (10.21) | **<0.001** |
| Sleep disorders | 98 (52.13) | 3061 (26.98) | **<0.001** |

COVID, those with SUD had a significantly higher prevalence of MHD than those without SUD (77.7% vs. 62.0%; $p<0.001$ , Table V). Furthermore, the onset of long COVID within the SUD population was associated with a substantially increased prevalence of major depressive disorder (43.1% vs. 30.4%), other anxiety disorders (50.5% vs. 36.8%), and sleep disorders (52.1% vs. 27.0%) (all $p<0.001$ , Table VI).

A multivariable logistic regression analysis quantified this risk, showing that specific SUDs dramatically increased the odds of a co-occurring MHD among long COVID patients (Table VII). The risk was more than doubled for Alcohol (AOR: 2.37), Opioid (AOR: 2.40), and Cannabis (AOR: 2.10) use disorders, and was highest for Cocaine Use Disorder (AOR: 2.82). In contrast, Tobacco Use Disorder was associated with a slightly reduced likelihood of a co-diagnosed MHD (AOR: 0.84), a finding that may warrant further study.

**TABLE VII:** Adjusted Odds Ratios (aOR) demonstrating the association between SUD and MHD among long-COVID patients.

| Exposure | AOR | 95% CI Lower | 95% CI Upper |
| --- | --- | --- | --- |
| Alcohol Use Disorder | 2.366 | 2.1068 | 2.6571 |
| Opioid Use Disorder | 2.3981 | 2.1026 | 2.7352 |
| Cannabis Use Disorder | 2.1002 | 1.7913 | 2.4623 |
| Cocaine Use Disorder | 2.8175 | 2.1782 | 3.6443 |
| Tobacco Use Disorder | 0.8353 | 0.7717 | 0.9042 |

Note. Outcome = Mental Health Disorder (MHD). All associations were statistically significant with P-value < 0.001.

### C) Prediction of In-Hospital Mortality

#### 1. Training Results: The training results are displayed in Table VIII

Standard classifiers trained on the original, imbalanced data achieved artificially high accuracy (>98%) but had negligible F1 scores and recall. This indicates a failure to identify the minority (death) class, rendering them clinically unusable.

**TABLE VIII:** The training results of death prediction task.

| Imbalance Strategy | Model | Accuracy | Precision | Recall | F1 | ROC AUC | PR AUC |
| --- | --- | --- | --- | --- | --- | --- | --- |
| None (Baseline) | RandomForest | 0.99 | 0.30 | 0.07 | 0.12 | 0.93 | 0.37 |
|  | XGBoost | 0.98 | 0.38 | 0.12 | 0.16 | 0.94 | 0.39 |
|  | LogisticRegression | 0.99 | 0.51 | 0.30 | 0.37 | 0.90 | 0.37 |
|  | Tab-Transformer | 0.99 | 0.57 | 0.44 | 0.50 | 0.72 | - |
| Algorithmic | BalancedRandomForest | 0.93 | 0.15 | 0.84 | 0.25 | 0.96 | 0.29 |
|  | EasyEnsemble | 0.88 | 0.10 | 0.89 | 0.17 | 0.95 | 0.29 |
| SMOTE | RandomForest | 0.99 | 0.49 | 0.34 | 0.39 | 0.96 | 0.39 |
|  | XGBoost | 0.98 | 0.46 | 0.48 | 0.46 | 0.93 | 0.35 |
|  | LogisticRegression | 0.95 | 0.18 | 0.64 | 0.28 | 0.86 | 0.31 |
| SMOTEENN | RandomForest | 0.98 | 0.45 | 0.49 | 0.46 | 0.95 | 0.39 |
|  | XGBoost | 0.98 | 0.36 | 0.57 | 0.44 | 0.94 | 0.42 |
|  | LogisticRegression | 0.95 | 0.16 | 0.64 | 0.25 | 0.88 | 0.29 |

Algorithmic methods like Balanced Random Forest (BRF) and Easy Ensemble (EE) achieved the highest recall (0.84 and 0.89, respectively), successfully flagging the majority of at-risk patients but at the cost of very low precision. Data resampling techniques provided a more practical balance. Notably, Logistic Regression combined with SMOTE (LR-SMOTE) achieved a strong recall of 0.64 while maintaining a more balanced F1-score. Given that minimizing false negatives is paramount for mortality prediction, LR-SMOTE was selected as the most promising candidate for fine-tuning.

On the original data, the Transformer achieved a validation F1-score of 0.38 and a recall of 0.27 for mortality prediction. The observed underperformance compared to simpler models is consistent with recent evidence that tree-based and regularized linear models often outperform Transformers on structured tabular data, which lacks the spatial or sequential patterns that Transformers are designed to leverage [18], [19]. The high dimensionality and relatively small number of positive cases in our dataset likely favored the strong inductive biases of conventional models over the data-hungry nature of deep learning architectures.

#### 2) Fine-tuned Model Performance

Based on our screening, we selected BRF and LR-SMOTE for final hyperparameter tuning and evaluation on the held-out test set.

The results (Table IX) reveal a clear performance trade-off. The BRF model achieved excellent recall (0.88) but at the cost of very low precision (0.14), indicating a high rate of false positives. This imbalance is visualized in its confusion matrix (Figure 2 Left).

**TABLE IX:** Fine-tune results of death prediction task.

| Model | Accuracy | F1 Score | Recall | Precision | ROC AUC | PR AUC |
| --- | --- | --- | --- | --- | --- | --- |
| BalancedRandomForest | 0.91 | 0.25 | 0.88 | 0.14 | 0.95 | 0.26 |
| LogisticRegression (SMOTE) | 0.94 | 0.36 | 0.93 | 0.22 | 0.95 | 0.27 |

**Fig. 2:**
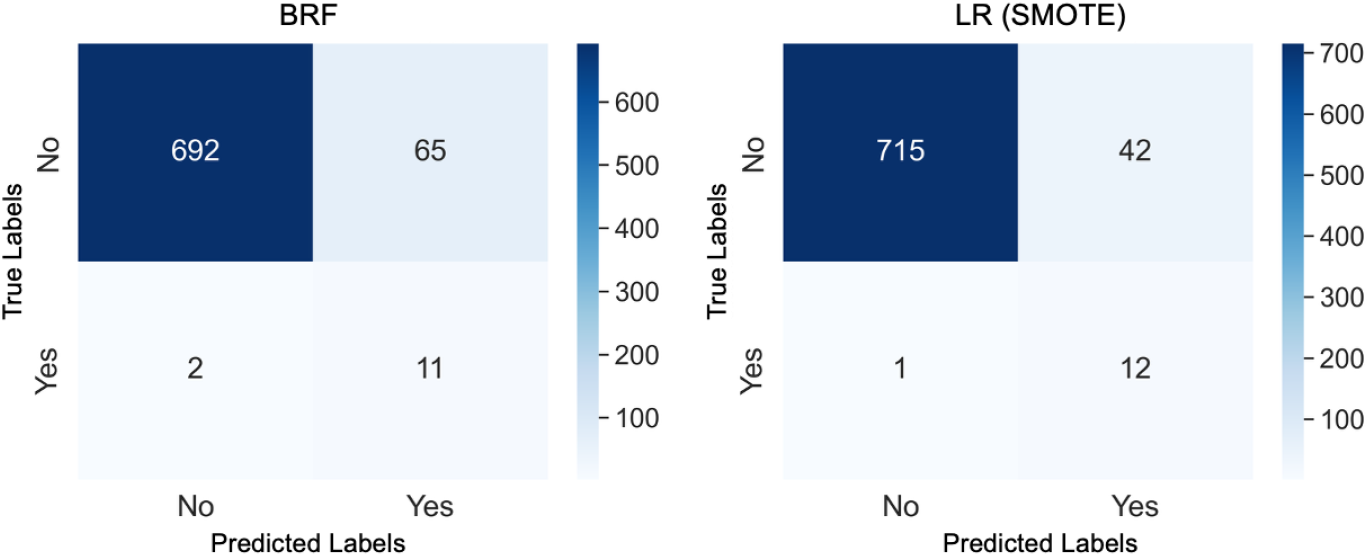
Confusion matrix of models for predicting death

In contrast, the LR-SMOTE model offered a more clinically actionable balance. It achieved an exceptional recall of 0.93 while improving precision to 0.22, resulting in a superior F1-score of 0.36. Its confusion matrix (Figure 2 Right) confirms this balance, showing it correctly identified all but one non-survivor while generating substantially fewer false alarms than BRF.

Therefore, we identified LR-SMOTE as the optimal model. It most effectively balances the competing clinical needs of maximizing the detection of at-risk patients (high recall) while minimizing the burden of false positives (higher precision).

#### 3) Feature Importance

The feature importance charts for the BRF and LR models reveal shared significance in features like “Lactic Acid”, “Age”, and “PO2 Arterial”, although their rankings vary between models (Figure 3). Elevated lactic acid, showed in both plots, is a predictor of mortality in trauma patients, linked to increased mortality in different critical illnesses [20], [21].

**Fig. 3:**
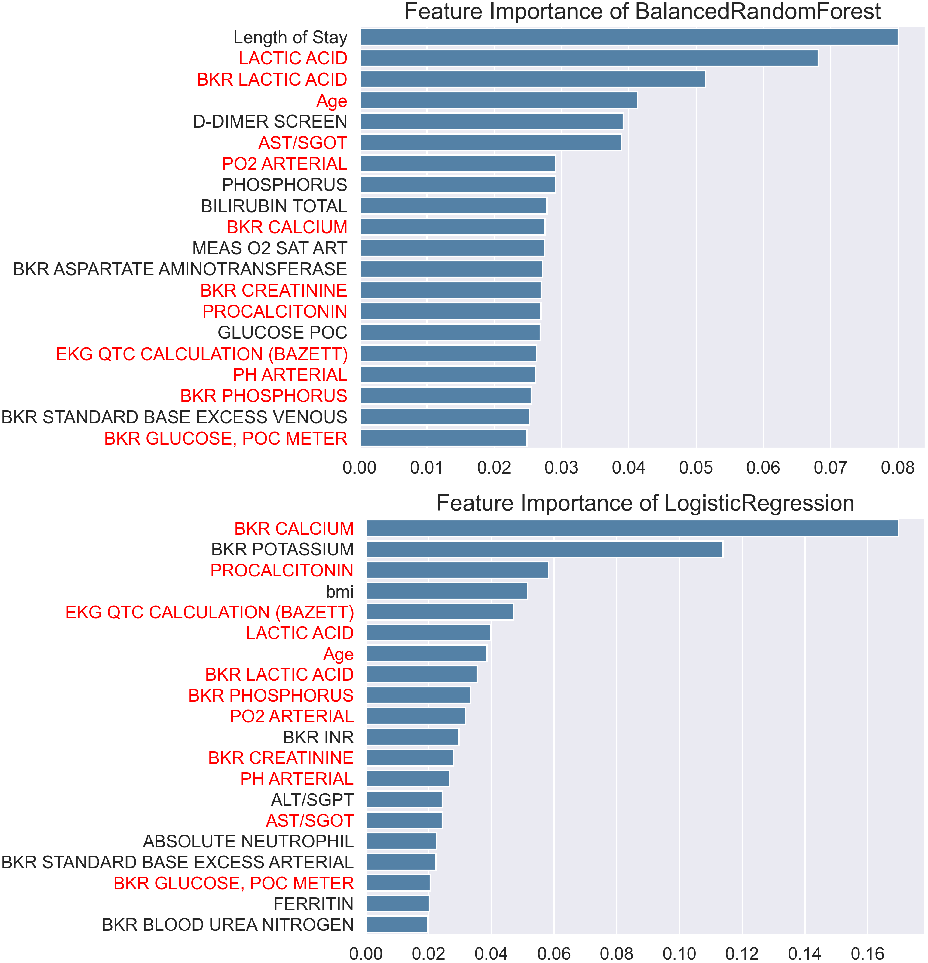
Feature importance related to in-hospital mortality calculated by models. The common significant features are highlighted in red.

“Length of Stay” is paramount for BRF, yet doesn’t feature as prominently for LR, highlighting the models’ distinct handling of data. “D-DIMER” was also considered an important feature. It has been correlated with thrombotic events and worse outcomes in COVID-19 patients [22], [23].

In terms of the results of LR, the model assigned greater importance to “BKR Calcium”, “BKR Potassium”, and “Procalcitonin” than BRF, illustrating different prioritizations in feature relevance for predicting outcomes.

### D. Prediction of Long COVID Development in the SUD Population

#### 1. Training Results

Initial experiments with standard classifiers on the original, imbalanced data demonstrated the profound difficulty of the task (Table X). Models like LR and XGBoost achieved deceptively high accuracy (97%) by simply classifying nearly all patients into the majority (non-long COVID) class. Their near-zero recall and F1 scores confirmed they had failed to learn any meaningful signal for predicting long COVID. In contrast, algorithmic approaches such as BRF and EE achieved the highest recall scores (0.77 and 0.71, respectively), proving their ability to identify at-risk patients, albeit with very low precision. Among the resampling techniques, LR combined with SMOTE and SMOTEENN was most effective, substantially boosting recall to 0.58 and 0.65.

Tab-Transformer, evaluated as an advanced baseline, performed similarly to the other standard classifiers on the raw data, achieving a recall of just 0.14 and an F1-score of 0.16. This underperformance is consistent with our mortality prediction results and reinforces that for high-dimensional, imbalanced tabular data, conventional models paired with robust sampling strategies often outperform deep learning architectures.

**TABLE X:** The training results of long COVID prediction task.

| Imbalance Strategy | Model | Accuracy | Precision | Recall | F1 | ROC AUC | PR AUC |
| --- | --- | --- | --- | --- | --- | --- | --- |
| None (Baseline) | RandomForest | 0.97 | 0.00 | 0.00 | 0.00 | 0.72 | 0.09 |
|  | XGBoost | 0.97 | 0.15 | 0.02 | 0.03 | 0.68 | 0.10 |
|  | LogisticRegression | 0.97 | 0.13 | 0.03 | 0.04 | 0.76 | 0.11 |
|  | Tab-Transformer | 0.96 | 0.20 | 0.14 | 0.16 | 0.56 | - |
| Algorithmic | BalancedRandomForest | 0.66 | 0.06 | 0.77 | 0.12 | 0.75 | 0.11 |
|  | EasyEnsemble | 0.66 | 0.06 | 0.71 | 0.11 | 0.71 | 0.11 |
| SMOTE | RandomForest | 0.96 | 0.11 | 0.05 | 0.06 | 0.76 | 0.09 |
|  | XGBoost | 0.95 | 0.09 | 0.06 | 0.07 | 0.73 | 0.09 |
|  | LogisticRegression | 0.76 | 0.07 | 0.58 | 0.12 | 0.73 | 0.10 |
| SMOTEENN | RandomForest | 0.94 | 0.10 | 0.13 | 0.11 | 0.76 | 0.08 |
|  | XGBoost | 0.93 | 0.10 | 0.19 | 0.13 | 0.76 | 0.09 |
|  | LogisticRegression | 0.72 | 0.06 | 0.65 | 0.12 | 0.73 | 0.11 |

#### 2) Fine-tuned Model Performance

For fine-tuned results, XGBoost with SMOTEENN achieved the highest overall precision (0.94) and a balanced F1 score (0.61), indicating robust general predictive power. However, to identify at-risk individuals, our proposed Custom Weighted Voting model demonstrated a stronger ability to capture positive cases, achieving the highest Recall (0.80) and PR AUC (0.21) among the evaluated methods (Table XI). This suggests a strategic optimization aimed at minimizing false negatives, which is crucial for a screening application where missing a case can have significant clinical consequences.

**TABLE XI:** Fine-tuning results of long COVID prediction task.

| Model | Accuracy | F1 Score | Recall | Precision | ROC AUC | PR AUC |
| --- | --- | --- | --- | --- | --- | --- |
| LogisticRegression (SMOTEENN) | 0.75 | 0.49 | 0.71 | 0.53 | 0.72 | 0.1 |
| XGBoost (SMOTEENN) | 0.94 | 0.61 | 0.65 | 0.59 | 0.84 | 0.18 |
| BalancedRandomForest | 0.71 | 0.48 | 0.78 | 0.54 | 0.86 | 0.19 |
| CustomWeightedVoting (Proposed) | 0.78 | 0.52 | 0.8 | 0.55 | 0.86 | 0.21 |

The confusion matrix (Figure 4) shows that the model correctly identified 23 long COVID cases (True Positive) and missed 4 (False Negative), achieving a high recall of 0.85 for the positive class. The high recall aligned with the 0.80 in the table, and PR AUC (0.21 from the table) underscores the model’s effectiveness as a sensitive screening tool. Although the low precision (0.11 from the matrix) indicates that a substantial proportion of positive predictions will be false alarms, this is often an acceptable trade-off in early-stage disease detection.

**Fig. 4:**
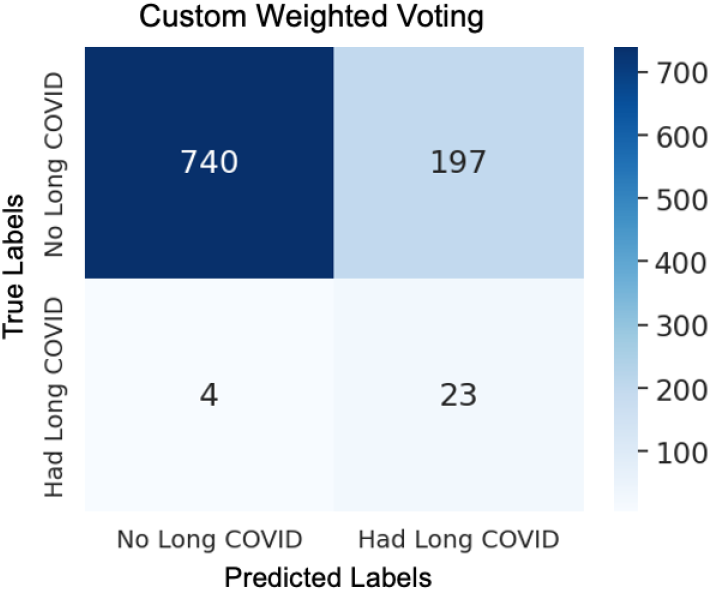
Confusion matrix of CustomWeightedVoting model for predicting long COVID.

#### 3) Feature Importance

Feature importance analysis of our well-performing models revealed a clinically coherent, multi-system signature for predicting long COVID, grounded in existing literature (Figure 5).

**Fig. 5:**
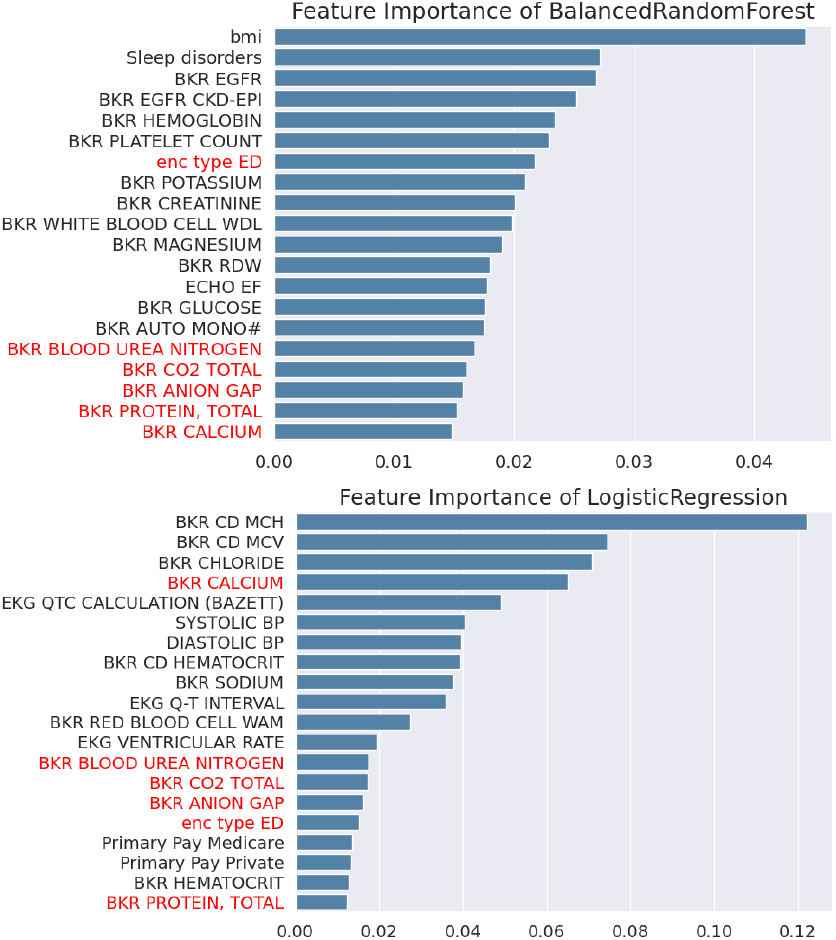
Feature importance calculated by models predicting long COVID.

The BRF model prioritized patient-level factors like Body Mass Index (BMI) and pre-existing Sleep Disorders. This aligns with strong evidence identifying obesity as a major risk factor, likely due to chronic inflammation and adipose tissue acting as a viral reservoir [24], [25]. The strong contribution of sleep disorders is also consistent with studies linking poor sleep quality to the development of long COVID, possibly through its effects on inflammation and mental health [26]– [28].

Both models also converged on the importance of laboratory markers reflecting renal and hematological dysfunction. The BRF model emphasized markers of kidney function, such as Estimated Glomerular Filtration Rate (EGFR) and Creatinine, a significant finding as declining renal function is a common long COVID sequela [29]. Concurrently, both models highlighted hematological dysregulation. The LR model prioritized red blood cell (RBC) indices like MCH and MCV, while the BRF focused on Hemoglobin and RDW. This shared emphasis points to persistent alterations in RBC health and oxygen-carrying capacity as a core feature of the long COVID phenotype [30], [31].

Finally, the models captured signals of both systemic distress and socioeconomic vulnerability. Predictive electrolytes like Chloride and Calcium likely reflect underlying renal dysfunction and inflammation [32], [33]. Crucially, the inclusion of encounter type (emergency department visit) and insurance status underscores the role of social determinants of health, confirming that cumulative social disadvantage is a significant risk factor for developing long COVID [34].

## V. Discussion

This study underscores the compounded vulnerabilities of patients with Substance Use Disorders (SUD) by developing robust machine learning models to predict distinct acute (in-hospital mortality) and chronic (long COVID) risks. Our primary contribution lies in demonstrating that imbalance-aware conventional ML models, specifically Logistic Regression with SMOTE (for mortality) and a custom weighted ensemble (for long COVID), can effectively stratify risk in this high-risk population. The mortality model achieved an exceptional 93% recall, indicating its potential for early identification of critically ill patients, while the long COVID model demonstrated an 80% recall, providing a crucial screening tool for prolonged illness. These findings highlight the power of computational approaches to address the critical need for proactive clinical decision support in managing the syndemic of COVID-19 and SUD.

The clinical utility of these models is significant. The mortality model could be integrated into a hospital’s Electronic Health Record (EHR) to flag high-risk patients upon admission, prompting closer monitoring and early intervention. Similarly, the long COVID model could be deployed as a screening tool at discharge, automatically referring at-risk individuals to specialized care and potentially mitigating the vicious cycle of chronic illness and substance use relapse.

Importantly, our investigation into deep learning architectures revealed that established tree-based and regularized linear models significantly outperformed the Tab-Transformer on this structured, imbalanced tabular EHR data. The reason is that conventional models possess stronger inductive biases, better suited for tabular data lacking the spatial or sequential structure that deep learning excels [18], [19]. The limited positive case counts in our dataset likely further challenged the data-hungry Transformer architecture. This reinforces the continued relevance of robust, interpretable traditional ML methods in clinical analytics.

We acknowledge several limitations, primarily stemming from the use of historical data from a single healthcare system. This may restrict the generalizability of our findings to different patient demographics and geographic areas. Furthermore, the models’ efficacy depends on the quality of available data and the specific clinical definitions used to identify SUD and long COVID, which may evolve over time.

In the future, the potential work can be:

### a) Longitudinal and Multi-Center Studies

Prospective studies are needed to validate our predictive models and track the long-term trajectory of mental and physical health in patients with SUD and long COVID. Expanding the data set to include various healthcare networks would significantly increase the generalizability of the model.

### b) Impact of Vaccination

While preliminary evidence suggests vaccination may reduce the risk of long COVID, its specific efficacy in SUD populations, who may have differing immune responses, remains unclear [35]. Future work should investigate the impact of vaccine timing, type, and dosage in this specific cohort, as booster vaccinations are critical for protection in patients with SUD [36].

### c) Advanced Machine Learning

Although our tab-transformer baseline did not outperform conventional models, future work could explore more advanced techniques. This includes multi-modal models that integrate structured EHR data with unstructured data like clinical notes and medical images, which may provide the rich, contextual information needed to further enhance predictive accuracy and clinical utility.

## VI. CONCLUSION

This research successfully developed and validated different machine learning models to predict in-hospital mortality and long COVID development among hospitalized COVID-19 patients with Substance Use Disorder (SUD). Our findings demonstrate that traditional machine learning approaches that are aware of imbalance, specifically Logistic Regression with SMOTE for mortality and a custom weighted ensemble for long COVID, can achieve clinically relevant predictive performance in this high-risk population. By providing robust predictive tools, this work offers a critical step toward proactive clinical decision support, enabling targeted interventions to mitigate both immediate mortality and long-term chronic disease in a population profoundly affected by the COVID-19 syndemic. The comparative evaluation also confirmed the continued strength of traditional ML methods over deep learning architectures in complex, imbalanced tabular EHR data, offering practical guidance for future clinical informatics research.

## Data Availability

All data produced in the present study are available upon reasonable request to the authors

## Acknowledgment

We gratefully acknowledge the support of the Northwestern Mutual Data Science Institute (NMDSI) for funding this project.

## References

[1] D. Benzano, F. Ornell, J. B. Schuch, F. Pechansky, A. O. Sordi, L. von Diemen, and F. H. P. Kessler, “Clinical vulnerability for severity and mortality by covid-19 among users of alcohol and other substances,” Psychiatry Research, vol. 300, p. 113915, 2021.

[2] Y. Sun, Y. Bao, T. Kosten, J. Strang, J. Shi, and L. Lu, “Challenges to opioid use disorders during covid-19,” The American journal on addictions, vol. 29, no. 3, p. 174, 2020.

[3] Q. Q. Wang, D. C. Kaelber, R. Xu, and N. D. Volkow, “Covid-19 risk and outcomes in patients with substance use disorders: analyses from electronic health records in the united states,” Molecular psychiatry, vol. 26, no. 1, pp. 30–39, 2021.

[4] D. C. Sanchez-Ramirez, K. Normand, Y. Zhaoyun, and R. Torres-Castro, “Long-term impact of covid-19: a systematic review of the literature and meta-analysis,” Biomedicines, vol. 9, no. 8, p. 900, 2021.

[5] A. D. Desai, M. Lavelle, B. C. Boursiquot, and E. Y. Wan, “Long-term complications of covid-19,” American Journal of Physiology-Cell Physiology, vol. 322, no. 1, pp. C1–C11, 2022.

[6] N. Panchal, R. Kamal, K. Orgera, C. Cox, R. Garfield, L. Hamel, and P. Chidambaram, “The implications of covid-19 for mental health and substance use,” Kaiser family foundation, vol. 21, pp. 1–16, 2020.

[7] L. Graig and K. Friedman, “Mental health and substance use disorders in the era of covid-19: The impact of the pandemic on communities of color: Proceedings of a workshop—in brief,” 2021.

[8] J. Wu, P. Annapureddy, P. Madiraju, S. I. Ahamed, J. Hernandez-Meier, C. Kostelac, and J. Dickson-Gomez, “Analysis and prediction of in-hospital risks among covid-19 patients with substance use disorder (sud),” in 2024 IEEE 48th Annual Computers, Software, and Applications Conference (COMPSAC). IEEE, 2024, pp. 833–842.

[9] A. R. Board, S. Kim, J. Park, L. Schieber, G. F. Miller, J. Pike, L. J. Cremer, and A. Asher, “Risk factors for covid-19 among persons with substance use disorder (pwsud) with hospital visits–united states, april 2020–december 2020,” Drug and Alcohol Dependence, vol. 232, p. 109297, 2022.

[10] C. C. Tam, S. Qiao, C. Garrett, R. Zhang, A. Aghaei, A. Aggarwal, A. H. Litwin, and X. Li, “Substance use, psychiatric symptoms, personal mastery, and social support among covid-19 long haulers: A compensatory model,” PLoS One, vol. 18, no. 8, p. e0289413, 2023.

[11] L. Naji, B. Dennis, R. L. Morgan, N. Sanger, A. Worster, J. Paul, L. Thabane, and Z. Samaan, “Investigating and addressing the immediate and long-term consequences of the covid-19 pandemic on patients with substance use disorders: a scoping review and evidence map protocol,” BMJ open, vol. 11, no. 9, p. e045946, 2021.

[12] F. Cascini, F. Santaroni, R. Lanzetti, G. Failla, A. Gentili, and W. Ricciardi, “Developing a data-driven approach in order to improve the safety and quality of patient care,” Frontiers in public health, vol. 9, p. 667819, 2021.

[13] E. Kriegova, M. Kudelka, M. Radvansky, and J. Gallo, “A theoretical model of health management using data-driven decision-making: the future of precision medicine and health,” Journal of Translational Medicine, vol. 19, no. 1, pp. 1–12, 2021.

[14] M. Purohit and P. Madiraju, “Predicting mental health disorders post long covid diagnosis using advanced machine learning techniques,” in 2023 IEEE International Conference on Big Data (BigData). IEEE, 2023, pp. 4954–4962.

[15] B. K. Patterson, J. Guevara-Coto, J. Mora, E. B. Francisco, R. Yogendra, R.A. Mora-Rodríguez, C. Beaty, G. Lemaster, G. Kaplan DO, A. Katz et al., “Long covid diagnostic with differentiation from chronic lyme disease using machine learning and cytokine hubs,” Scientific Reports, vol. 14, no. 1, p. 19743, 2024.

[16] X. Huang, A. Khetan, M. Cvitkovic, and Z. Karnin, “Tabtransformer: Tabular data modeling using contextual embeddings,” arXiv preprint 2012.06678, 2020.

[17] V. Kumar, G. S. Lalotra, and R. K. Kumar, “Improving performance of classifiers for diagnosis of critical diseases to prevent covid risk,” Computers and Electrical Engineering, vol. 102, p. 108236, 2022.

[18] R. Shwartz-Ziv and A. Armon, “Tabular data: Deep learning is not all you need,” Information Fusion, vol. 81, pp. 84–90, 2022.

[19] L. Grinsztajn, E. Oyallon, and G. Varoquaux, “Why do tree-based models still outperform deep learning on typical tabular data?” Advances in neural information processing systems, vol. 35, pp. 507–520, 2022.

[20] L. W. Andersen, J. Mackenhauer, J. C. Roberts, K. M. Berg, M. N. Cocchi, and M. W. Donnino, “Etiology and therapeutic approach to elevated lactate levels,” in Mayo Clinic Proceedings, vol. 88, no. 10. Elsevier, 2013, pp. 1127–1140.

[21] F. O. Giuste, L. He, P. Lais, W. Shi, Y. Zhu, A. Hornback, C. Tsai, M. Isgut, B. Anderson, and M. D. Wang, “Early and fair covid-19 outcome risk assessment using robust feature selection,” Scientific Reports, vol. 13, no. 1, p. 18981, 2023.

[22] L. Zhang, X. Yan, Q. Fan, H. Liu, X. Liu, Z. Liu, and Z. Zhang, “Ddimer levels on admission to predict in-hospital mortality in patients with covid-19,” Journal of thrombosis and haemostasis, vol. 18, no. 6, pp. 1324–1329, 2020.

[23] T. W. Tulu, T. K. Wan, C. L. Chan, C. H. Wu, P. Y. M. Woo, C. Z. S. Tseng, A. Vodencarevic, C. Menni, and K. H. K. Chan, “Machine learning-based prediction of covid-19 mortality using immunological and metabolic biomarkers,” BMC Digital Health, vol. 1, no. 1, p. 6, 2023.

[24] L. Vimercati, L. De Maria, M. Quarato, A. Caputi, L. Gesualdo, G. Migliore, D. Cavone, S. Sponselli, A. Pipoli, F. Inchingolo et al., “Association between long covid and overweight/obesity,” Journal of Clinical Medicine, vol. 10, no. 18, p. 4143, 2021.

[25] J. Fernández-García, A. Romero-Secin, and M. Rubín-García, “Association between obesity and long-covid: A narrative review,” Semergen, vol. 51, no. 3, p. 102390.

[26] W. Tański, A. Tomasiewicz, and B. Jankowska-Polańska, “Sleep disturbances as a consequence of long covid-19: Insights from actigraphy and clinimetric examinations—an uncontrolled prospective observational pilot study,” Journal of clinical medicine, vol. 13, no. 3, p. 839, 2024.

[27] H. Shao, H. Chen, K. Xu, Q. Gan, M. Chen, Y. Zhao, S. Yu, Y. K. Li, L. Chen, and B. Cai, “Investigating the associations between covid-19, long covid, and sleep disturbances: cross-sectional study,” JMIR Public Health and Surveillance, vol. 10, p. e53522, 2024.

[28] P. Xue, I. Merikanto, E. A. Delale, A. Bjelajac, J. Yordanova, R. N. Chan, M. Korman, S. A. Mota-Rolim, A.-M. Landtblom, K. Matsui et al., “Associations between obesity, a composite risk score for probable long covid, and sleep problems in sars-cov-2 vaccinated individuals,” International journal of obesity, vol. 48, no. 9, pp. 1300–1306, 2024.

[29] S. Copur, M. Berkkan, C. Basile, K. Tuttle, and M. Kanbay, “Post-acute covid-19 syndrome and kidney diseases: what do we know?” Journal of nephrology, vol. 35, no. 3, pp. 795–805, 2022.

[30] G. C. Lechuga, C. M. Morel, and S. G. De-Simone, “Hematological alterations associated with long covid-19,” Frontiers in physiology, vol. 14, p. 1203472, 2023.

[31] J. Bros, L. Ibershoff, E. Zollmann, J. Zacher, F. Tomschi, H.-G. Predel, W. Bloch, and M. Grau, “Changes in hematological and hemorheological parameters following mild covid-19: a 4-month follow-up study,” Hematology reports, vol. 15, no. 4, pp. 543–554, 2023.

[32] M. S. Mohamed, E. M. Negm, M. H. Zahran, M. M. Magdy, A. A. Mohammed, D. A. Ibrahim, A. E. Tawfik, and T. H. Hassan, “Electrolyte profile in covid-19 patients: insights into outcomes,” The Egyptian Journal of Bronchology, vol. 17, no. 1, p. 48, 2023.

[33] H. Song, A. Chia, B. Tan, C. Teo, V. Lim, H. Chua, M. Samuel, and A. Kee, “Electrolyte imbalances as poor prognostic markers in covid-19: a systemic review and meta-analysis,” Journal of endocrinological investigation, vol. 46, no. 2, pp. 235–259, 2023.

[34] J. Xiang, H. Zheng, Y. Cai, S. Chen, Y. Wang, and R. Chen, “Cumulative social disadvantage and its impact on long covid: insights from a us national survey,” BMC medicine, vol. 23, no. 1, p. 207, 2025.

[35] O. Byambasuren, P. Stehlik, J. Clark, K. Alcorn, and P. Glasziou, “Effect of covid-19 vaccination on long covid: systematic review,” BMJ medicine, vol. 2, no. 1, p. e000385, 2023.

[36] Y. Huang, “Covid-19 vaccine effectiveness in people with substance use disorder,” The Lancet Psychiatry, vol. 10, no. 6, pp. 372–373, 2023.

